# Age-Specific Rates of Onset of Cannabis Use in Mexico

**DOI:** 10.1101/2020.11.30.20241463

**Authors:** Mauricio López-Méndez, Angélica Ospina-Escobar, Rowan Iskandar, Fernando Alarid-Escudero

## Abstract

**Background:** Over the previous two decades, the prevalence of cannabis use has risen among the population in Mexico.

**Aims:** To estimate the sex- and age-specific rates of onset of cannabis use over time.

**Design:** Time-to-event flexible parametric models with spline specifications of the hazard function. Stratified analysis by sex, and control for temporal trends by year of data collection or decennial birth-cohort.

**Setting:** Mexico.

**Participants:** Pooled sample of 141,342 respondents aged between 12 and 65 years from five nationally representative cross-sectional surveys, the Mexican National Surveys of Addictions (1998, 2002, 2008, 2012) and the Mexican National Survey on Drugs, Alcohol, and Tobacco Consumption (2016).

**Measurements:** We estimated age-specific rates of onset of cannabis as the conditional rate of consuming cannabis for the first time at a specific age.

**Findings:** Age-specific rates of onset of cannabis use per 1,000 individuals increased over time for both females and males. Peak rates per 1,000 ranged from 0.935 (95%CI= [0.754,1.140]) in 1998, to 5.390 (95%CI= [4.910,5.960]) in 2016 for females; and from 7.510 (95%CI= [5.516, 10.355]) in 1998, to 26.100 (95%CI= [23.162,30.169]) in 2016 for males. Across decennial birth-cohorts, peak rates of onset of cannabis use per 1,000 individuals for females ranged from 0.342 (95%CI= [0.127,0.898]) for those born in the 1930s, to 14.600 (95%CI= [13.200,16.100]) for those born in the 1990s; and for males, from 4.900 (95%CI= [0.768, 7.947]) for those born in the 1930s, to 38.700 (95%CI= [32.553,66.341]) for those born in the 1990s.

**Conclusion:** Rates of onset of cannabis use for males are higher than for females; however, the change across recent cohorts of the rates of onset has increased at a faster rate among females. Our findings can inform and improve the implementation of policies around cannabis use by identifying subpopulations by age, sex, and birth-cohort that are at the highest risk of initiating cannabis consumption.

## Introduction

In Mexico, cannabis is the most commonly used illicit substance among the population between 12 and 65 years of age. In 2016, the Mexican National Institute of Public Health (INSP, in Spanish) reported that lifetime cannabis use prevalence was 8.6%, followed by cocaine (3.5%) and inhalants (1.1%) [29]. Cannabis use prevalence for males (14%) was higher than for females (3.7%), but for adolescents (12-17 years of age), the gap between males and females had decreased over the previous two decades. Among male adolescents, cannabis use prevalence increased from 2.1% in 2002 to 5.8% in 2016, and from 0.2% in 2002 to 4.8% in 2016 among female adolescents [29].

While the body of evidence on the potential harms and benefits associated with cannabis use is mixed [21, 33, 5, 6], there is substantial evidence that early age of onset is an important and understudied factor associated with elevated rates of harms and injuries and long-term consequences. For example, reduced odds of high-school completion and degree attainment, and higher odds of developing mental disorders, respiratory diseases, impaired cognitive function, and substance use disorders in adulthood; that could lead to increased morbidity and mortality [4, 21, 33, 4, 7, 8, 31, 34, 35, 43].

Despite the increasing prevalence of use of cannabis in Mexico and their potential downstream consequences, there is a lack of evidence on the rates at which males and females initiate cannabis use, at what ages, and how these rates have changed over time. Authorities have collected data on illicit drug use in the country since 1998; however, there are currently no estimates on incidence measures of onset of cannabis use or other illicit substances at the national level (i.e., rates of onset) and for different sub-populations (i.e., sex- and age-specific).

The lack of evidence on the incidence of initiation limits the assessment and projections of national cannabis use trends, studies of economic and health burden, the design and evaluation of harm-reduction and prevention interventions, and future regulation in the country [19]. Specifically, sex- and age-specific incidence measures may facilitate the focalization of resources and prevention and harm-reduction interventions by identifying sub-populations at highest risk to initiate consumption and consequently may increase their efficacy. Incidence rates are more informative and reliable measures than prevalence estimates since they can account for truncation and censoring of responses [36]. These measures are also integral components of epidemiologic and mathematical policy models that aim to characterize the progression in time of patterns of use among the population [11, 17, 18], particularly in situations where the equivalent to life-tables are not available [37].

These gaps in knowledge have become increasingly important given the current debate in Mexico on the regulation of adult use of cannabis [39,40,41] and the gradual departure from policies centered on prohibition in the region of North America [32, 38], (e.g., Canada and 11 states in the United States of America). As the experience in other regions suggest, [41,42,43] there is an increasing need for evidence-based arguments to inform the design and implementation of wide-scale policies and regulations around use of cannabis [5,21].

Based on the existing prevalence trends in the country, we hypothesize that rates of onset of cannabis use have increased along with prevalence but that they differ by sex and their growth over time. In this study, we estimate the rate of onset of cannabis use among the Mexican population using flexible parametric time-to-event models. Specifically, we estimate the temporal progression across periods and birth-cohorts of the sex- and age-specific rates of onset of cannabis use. The paper is organized as follows: In the first section, we provide a description of the surveys and the responses used to estimate the epidemiological measures of interest. We then describe the statistical methods to estimate the age- and sex-specific rates of cannabis initiation. The main results of our analysis are presented in the next section. Lastly, we discuss the potential limitations of our analysis and highlight the implications of our results for future research and the drug policy around adult use of cannabis in Mexico.

## Methods

### Samples and participants

We use data on responses obtained from five cross-sectional waves of a probabilistic, multistage, stratified, nationally representative surveys: 1) the National Survey of Addictions for years 1998, 2002, 2008, 2011 [27, 28, 30]; and 2) the National Survey of Alcohol, Tobacco, and Drugs Consumption for the year 2016 [12]. In 2016, the new name for the survey was used to mark the difference between problematic and non-problematic drug use [12]. In total, we used a pooled sample of 141,342 interviews obtained from individuals between 12-65 years of age (mean age of 33 years), of which 44.6%(n=61,658) are male, and 56.4% (n=79,684) are female. The pooled sample covers a timespan of 18 calendar years (1998-2016) and captures individuals born across seven decennial birth-cohorts (1930-1990). Figure 1, shows the range of birth-years included in the pooled sample, representing the range of birth-years captured by each individual survey, and Table 1 provides additional sample characteristics.

**Table 1.**
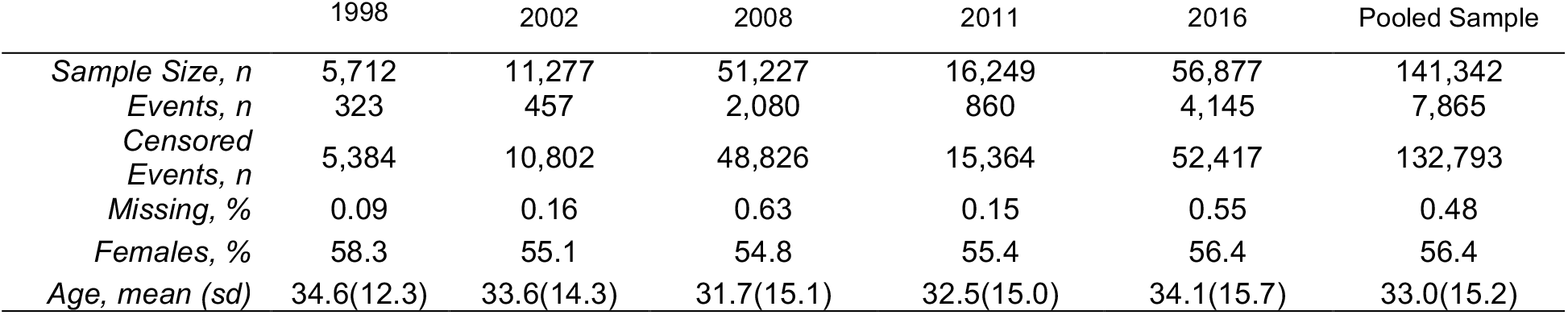
Sample Characteristics

**Figure 1.**
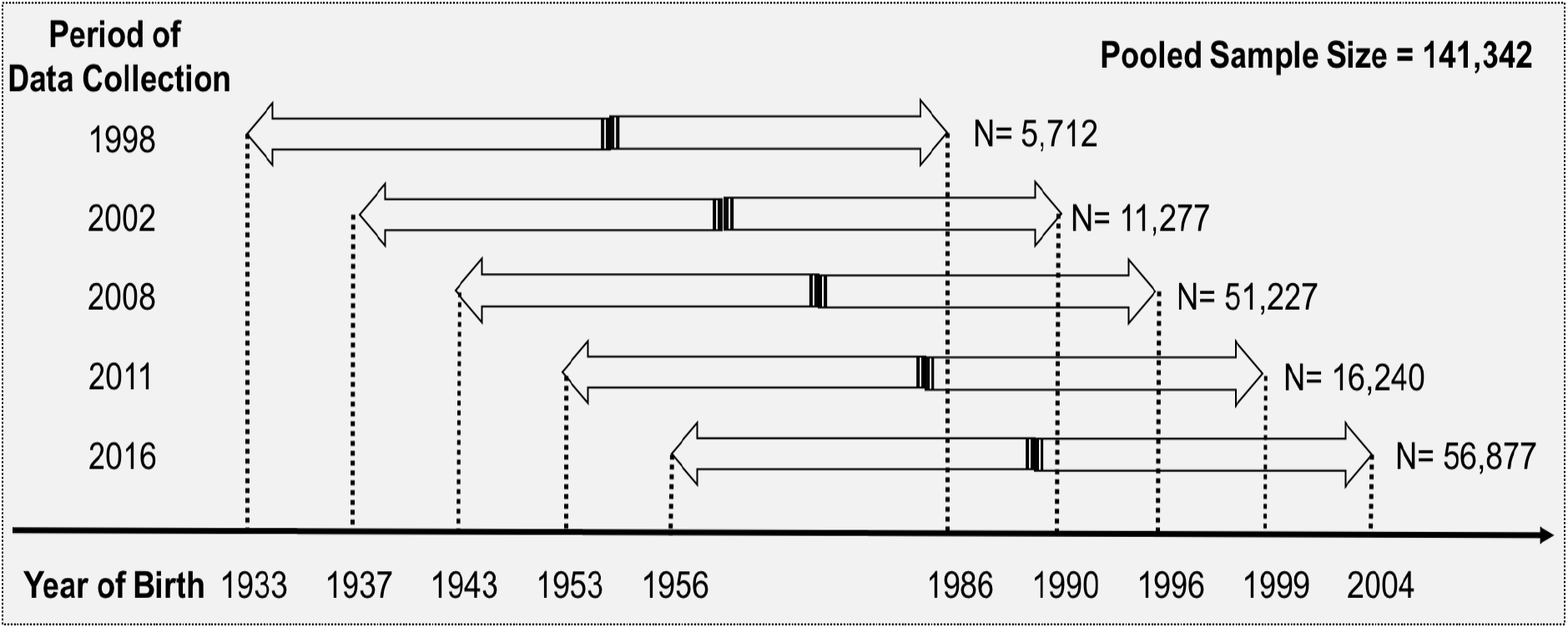
Range of Birth-Years included in the Pooled Sample

### Measures

We extracted self-reported responses for the following questions and items: “(…) ever used cannabis” (yes/no), “(…) use of cannabis in the last month” (yes/no), “(…) use of cannabis in the last 12 months” (yes/no)), “(…) age of first use of cannabis” (years of age), year of birth (calendar year), age (at time of the survey, years), and biological sex (male/female). We used the age of first use of cannabis as a measure of initiation because of its previously demonstrated reliability [15]. The age of first use represents the time elapsed in actuarial years since the birth of an individual until the age before cannabis use initiation. Within a time-to-event framework, the age of first use of cannabis was formulated as a time-to-event random variable. Accordingly, we used the responses on the age of first use to compute the age-specific rates of onset of cannabis use retrospectively. Age-specific rates of onset are the instantaneous and conditional rates of consuming cannabis at a specific age, given that it has not happened previously. This measure is also known as the “hazard rate” or “failure rate” in epidemiology, demography, and survival analysis [16].

We also estimated the sex-specific mean age of onset among those who reported ever having consumed cannabis across periods and birth cohorts in the sample. Mean age of onset among those who consumed cannabis is a statistic that represents the average age to initiate cannabis consumption among those who will consume cannabis in their lifetimes.

### Analysis

The mean age of onset among those who consumed cannabis was estimated retrospectively using the non-parametric Kaplan-Meier estimates among a subset of the data including only those observations that eventually consumed cannabis. The computation was done by calculating the area under the Kaplan-Meier curve among the life-time cannabis users.

To estimate the age-specific rates of onset of cannabis use, we fitted flexible-parametric time-to-event models stratified by sex (female/male). For each stratum, we modeled the baseline log-cumulative rates of onset or log-cumulative odds of onset as a natural cubic spline [26, 9]. The choice between the scale of the dependent variable (log cumulative rates of onset vs log cumulative odds of onset) was based on the overall fit of the models (i.e. AIC). The hazard rates of onset of cannabis were recovered from the log-cumulative rates of onset/odds predicted by the best fit models.

We used log-rank tests to assess whether the distribution of failure-times (i.e. age of onset of cannabis use) differed by sex and period, and by sex and decennial birth-cohort. We estimated the temporal trends on the rates/odds of onset by adjusting models that include either period of data collection (1998, 2002, 2008, 2011, 2016) or decennial birth-cohort (1930-1990), using orthogonal polynomial terms that act additively on the log-cumulative rates/odds of onset. To test the hypothesis of non-linear growth of the rates of onset across periods and cohorts, we compared different degrees of the orthogonal polynomial models in the temporal trends (i.e., linear, quadratic, or cubic). To compare the trends across periods and cohorts between males and females we plot the trajectories of the baseline log-cumulative rates of onset/odds of onset predicted by the best fit models for males and for females. The trajectories represent the additive shifts in the baseline log-cumulative rates of onset/odds across cohorts and periods. We also compare estimates of the trend coefficients of models with the same specification of temporal trend (i.e. linear vs linear, quadratic vs quadratic, or cubic vs cubic) to contrast the growth across periods and cohorts of the baseline log-cumulative rates of onset/odds for females and males based on the same structural temporal trend specification. Finally, we compared models by varying the number of internal knots on the natural cubic spline of the baseline cumulative rates and odds of onset. We selected the model specification with the lowest Akaike Information Criterion (AIC) [13, 26] for each stratum and temporal trend (period of data collection, decennial birth-cohort).

To estimate the age-specific rates of onset, we accounted for both “right” (data collected in 2002,208,2011, and 2016) and “interval” censoring of the data (data collected in 1998) under the assumption of independent (“non-informative”) censoring [16]. In the case of the computation of the mean age of onset among those who consumed cannabis, it was not possible to account for censoring since the sample was restricted to those who reported having consumed cannabis. We computed 95% confidence intervals of the parameters of the flexible-parametric models via empirical bootstrap using 300 replications of the original dataset for each sub-population [26]. A detailed characterization of these models as well as their mathematical formulation can be found in the Appendix (see supplemental material). The statistical analysis and modelling were conducted in R version 3.6.1 [25]. To fit the flexible parametric models of the age-specific rates of onset, we used the “flexsurv” R-package [13].

## Results

### Mean Age of Onset Among Life-time Cannabis Users

Table 2 shows the progression of the mean age of onset among those who consumed cannabis (“life-time cannabis users”) by sex, periods, and decennial birth cohorts (see Table 2, columns 2 and 3). Across periods, the mean age of onset among female lifetime cannabis users fluctuated between 20.6 (SE=0.940) years in 1998 and 17.7 (SE=0.185) years in 2016. Among male lifetime users, the mean age of onset decreased consistently from 19.3 (SE=0.316) years in 1998 to 17.4 (SE=0.090) years in 2016. Across cohorts, the mean age of onset among female lifetime users reached its peak at 24.3 (SE=1.689) years for those born in the 1950’s and consistently decreased to 15.7 (SE=0.091) years for those born in the 1990s. The mean age of onset among male lifetime users decreased consistently from 24.6 (SE=3.485) years for those born in the 1930s to 15.5 (SE=0.0651) years for those born in the 1990s.

**Table 2.** Mean Age of Onset and Peak Rate of Onset per 1,000 by Sex, Period and Decennial Birth Cohort

|  |  | Conditional Mean Age of Onset*<br>(SE) |  | Peak Rate of Onset per 1,000***<br>(Bootstrap 95%CI) |  |
| --- | --- | --- | --- | --- | --- |
|  |  | Female | Male | Female | Male |
| Period |  |  |  |  |  |
|  | 1998 | 20.600<br>(0.940) | 19.300<br>(0.317) | 0.935<br>(0.754,1.140) | 7.510<br>(5.516,10.355) |
|  | 2002 | 14.400<br>(0.849) | 13.400<br>(0.259) | 1.380<br>(1.190,1.620) | 15.200<br>(12.046,18.924) |
|  | 2008 | 17.500<br>(0.228) | 17.900<br>(0.124) | 1.380<br>(1.180,1.600) | 15.210<br>(14.246,19.497) |
|  | 2011 | 18.300<br>(0.505) | 17.900<br>(0.174) | 3.320<br>(3.030,3.650) | 16.100<br>(14.158,19.168) |
|  | 2016 | 17.700<br>(0.185) | 17.400<br>(0.090) | 5.390<br>(4.910,5.960) | 26.100<br>(23.162,30.169) |
| Decennial Cohort |  |  |  |  |  |
|  | 1930 | 17.00<br>(-)** | 24.600<br>(3.484) | 0.342<br>(0.127,0.898) | 4.900<br>(0.768, 7.947) |
|  | 1940 | 21.600<br>(2.091) | 23.600<br>(1.059) | 0.529<br>(0.359,0.770) | 8.270<br>(4.407,11.292) |
|  | 1950 | 24.300<br>(1.689) | 20.200<br>(0.335) | 0.656<br>(0.531,0.799) | 12.300<br>(9.900,16.109) |
|  | 1960 | 21.500<br>(0.704) | 18.300<br>(0.181) | 0.815<br>(0.700,0.958) | 15.500<br>(14.018,21.864) |
|  | 1970 | 19.700<br>(0.482) | 17.800<br>(0.124) | 1.270<br>(1.100,1.420) | 18.790<br>(17.407,29.284) |
|  | 1980 | 18.300<br>(0.251) | 17.100<br>(0.104) | 3.080<br>(3.271,3.480) | 28.500<br>(22.275,40.746) |
|  | 1990 | 15.700<br>(0.091) | 15.500<br>(0.065) | 14.600<br>(13.200,16.100) | 38.700<br>(32.553,66.341) |

\*The conditional mean age of onset is computed as the area under the Kaplan-Meier survival curve conditional on having experienced the event (i.e. having consumed cannabis).
\*\*The sample size for females in the 1930's was < 3 obs., estimate of the standard error is not reported.
\*\*\* Peak Rates of Onset per 1,000 were computed as the maximum rates of onset predicted by the flexible time to event models for each subpopulation (i.e. Period/Cohort and Sex).

### Log-rank-test

We found significant difference in the rates of onset by sex and period of data collection (Chi-squared = 387, d.f = 7, P-Value <0.000), and by sex and decennial birth-cohort (Chi-squared = 988, d.f = 12, P-Value <0.000).

### Age-specific Rates of Onset of Cannabis Use

Table 2 also shows the age-specific peak rates (i.e., the highest rate) of onset per 1,000 of cannabis use by sex, periods, and decennial birth cohorts (see Table 2, columns 4 and 5). In 1998, the peak rate of onset per 1,000 for females was 0.935 (95%CI= [0.754,1.140]), and increased to 5.390 (95%CI= [4.910,5.960]) in 2016. For males, in 1998 the peak rate of onset per 1,000 was 7.510 (95%CI= [6.655, 30.711]), and increased to 26.1 (95%CI= [24.160,44.177]) in 2016. Figure 2 shows the corresponding sex- and age-specific rates of onset per 1,000 by period of data collection.

**Figure 2.**
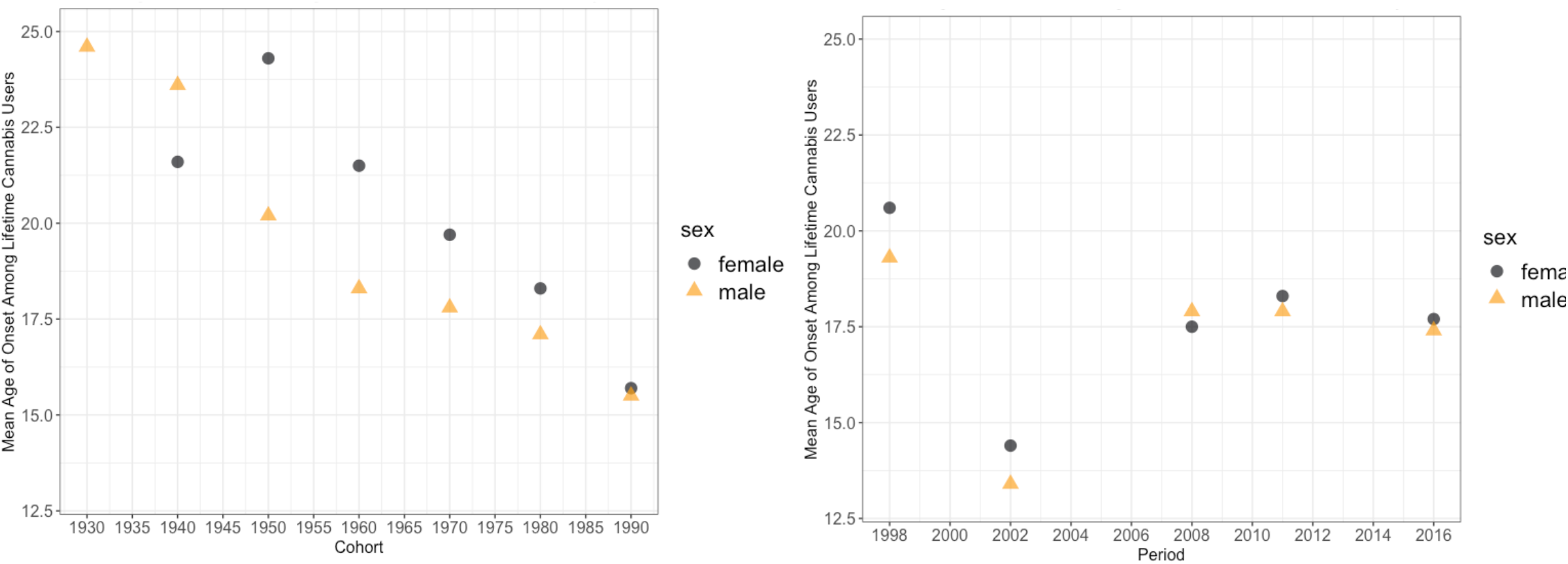
Mean Age of Onset Among Lifetime Cannabis Users, by Sex and Cohort/Period

For females born in the decade of 1930, the peak rate of onset per 1,000 individuals was 0.342 (95%CI= [0.127,0.898]), and increased to 14.600 (95%CI= [13.200,16.100]) for females born in the decade of 1990. For males born in the decade of 1930, the peak rate of onset per 1,000 individuals was 4.900 (95%CI= [4.498, 8.426]), and increased to 38.700 (95%CI= [34.610,62.660]) for males born in the decade of 1990. Figure 3 shows the corresponding sex- and age-specific rates of onset per 1,000 by decennial cohort.

**Figure 3.**
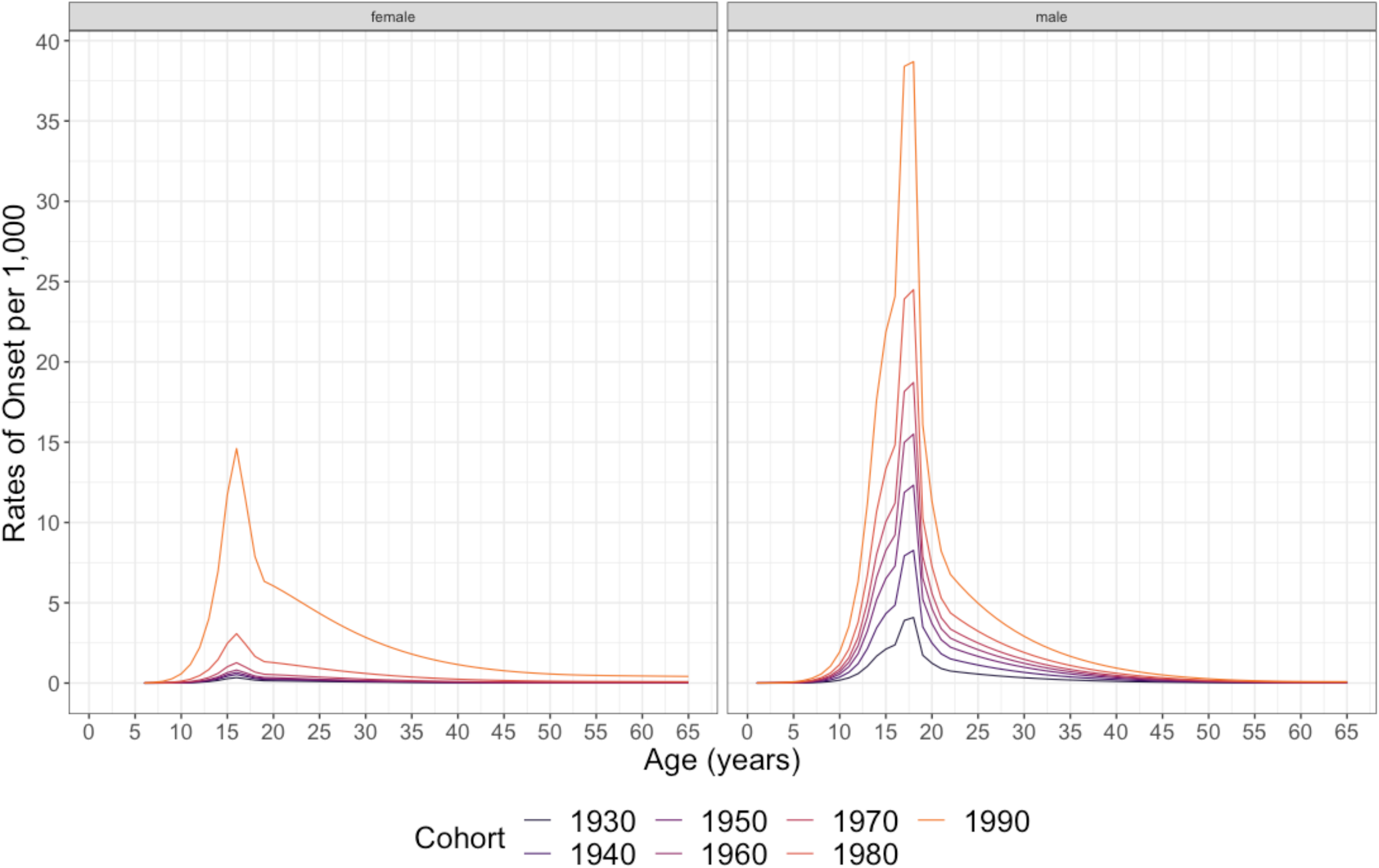
Age-Specific Rates of Onset of Cannabis Use per 1,000 by Sex and Cohort

**Figure 4.**
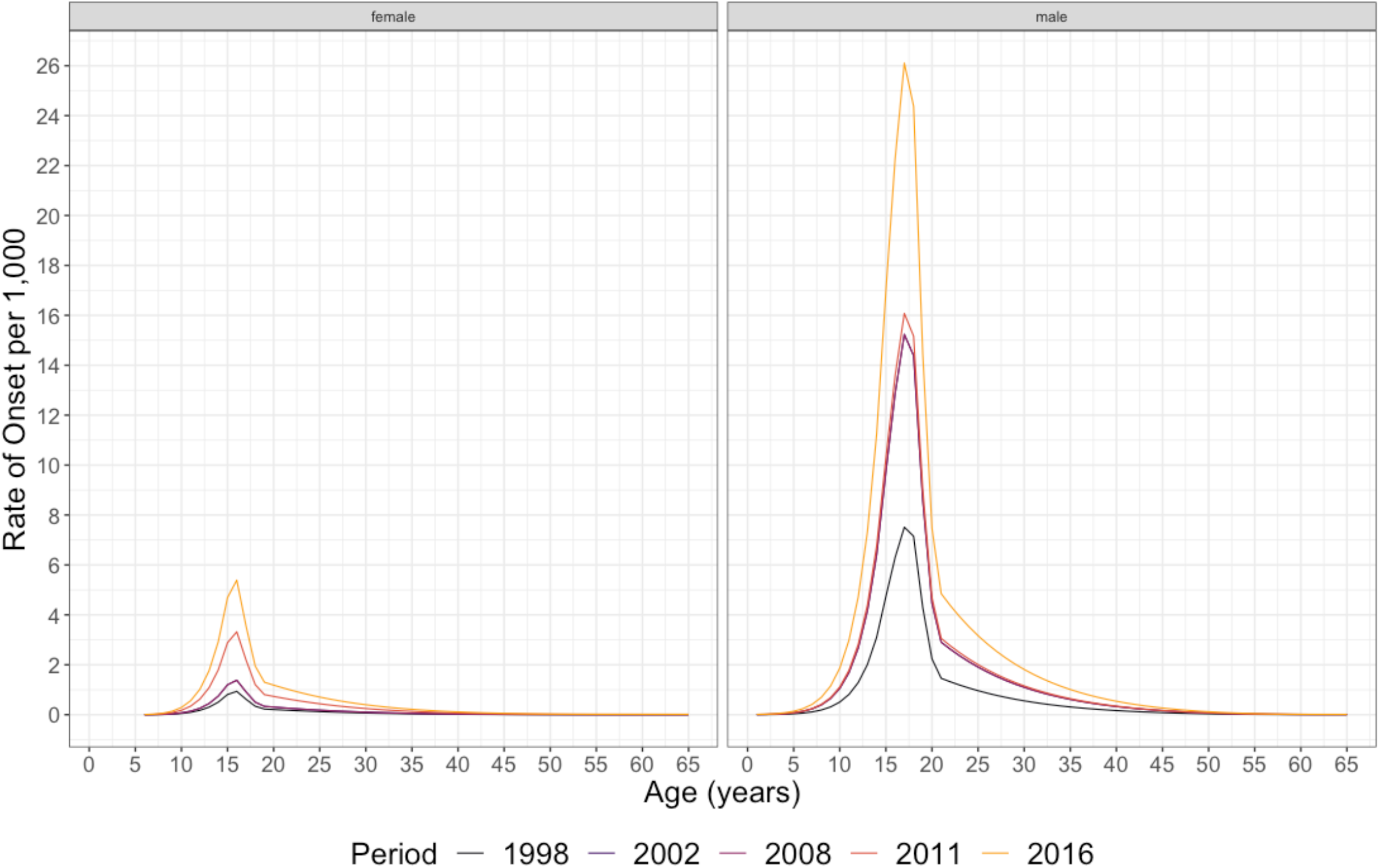
Age-Specific Rates of Onset of Cannabis Use per 1,000 by Sex and Period

### Temporal Trends of the Rates of Onset of Cannabis Use

Table 3 shows the estimates of the coefficients of the temporal trend terms for each of the best-fit models by sex. The rates of onset of cannabis use increased across periods and cohorts: however, the growth trajectories in the rates of onset across periods and cohorts differ by sex.

**Table 3.**
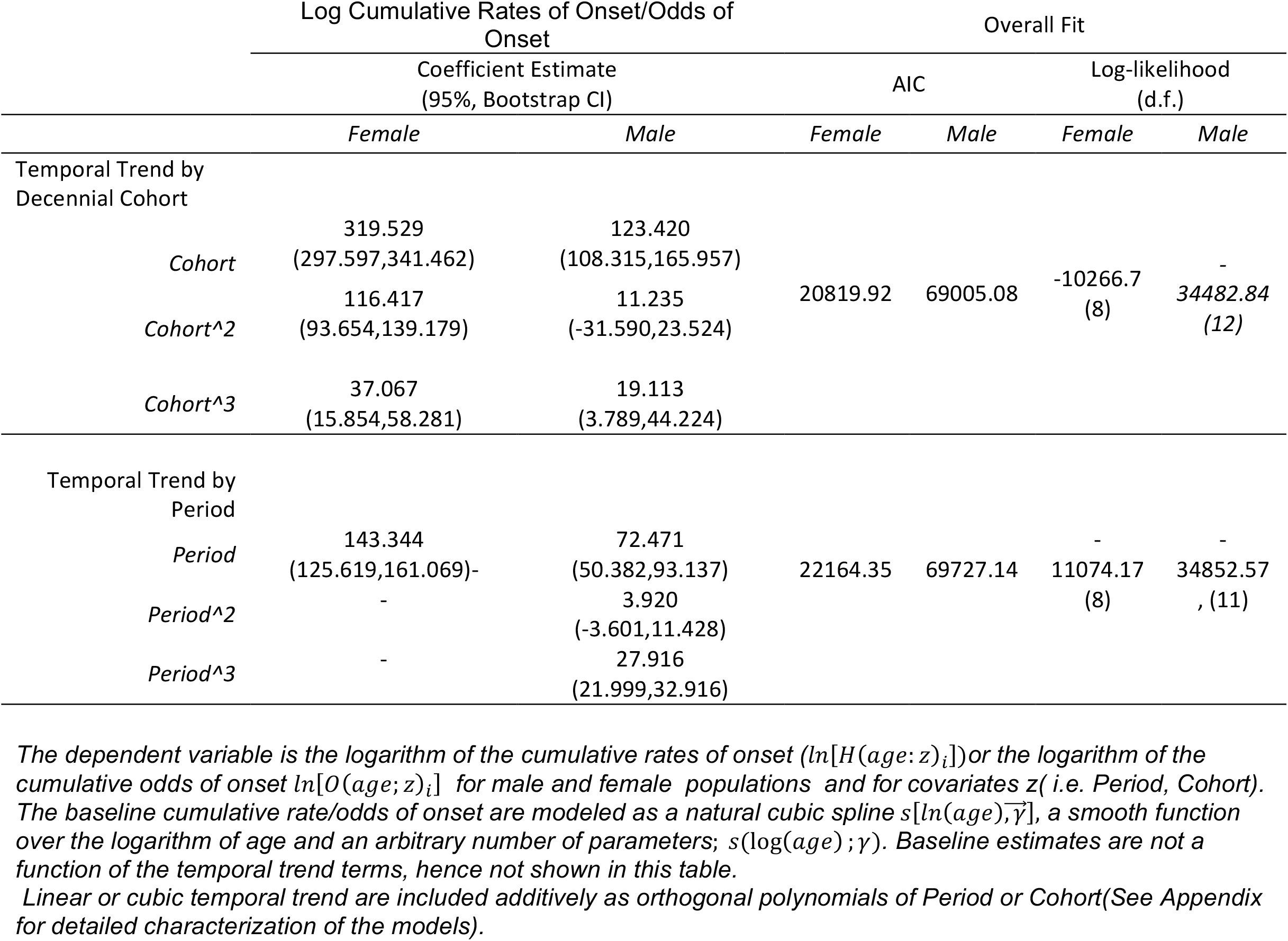
Best fit Models Results on Temporal Trend Coefficients on the Log Cumulative Rates of Onset/Odds of Onset

Figure 5 shows the temporal trend across cohorts predicted by the additive orthogonal polynomial trend terms by sex, representing the different trajectories across cohorts that the baseline log-cumulative rates of onset/odds of onset follow for females and males. Across decennial birth-cohorts, the temporal trend of the rates of onset of cannabis use among females and males were best described by a cubic trend, but for females, the trajectory across cohorts after the 1970s was much steeper than for males, this is also illustrated in the coefficient estimates of the temporal trend terms for females which are of larger magnitude than for males.

**Figure 5.**
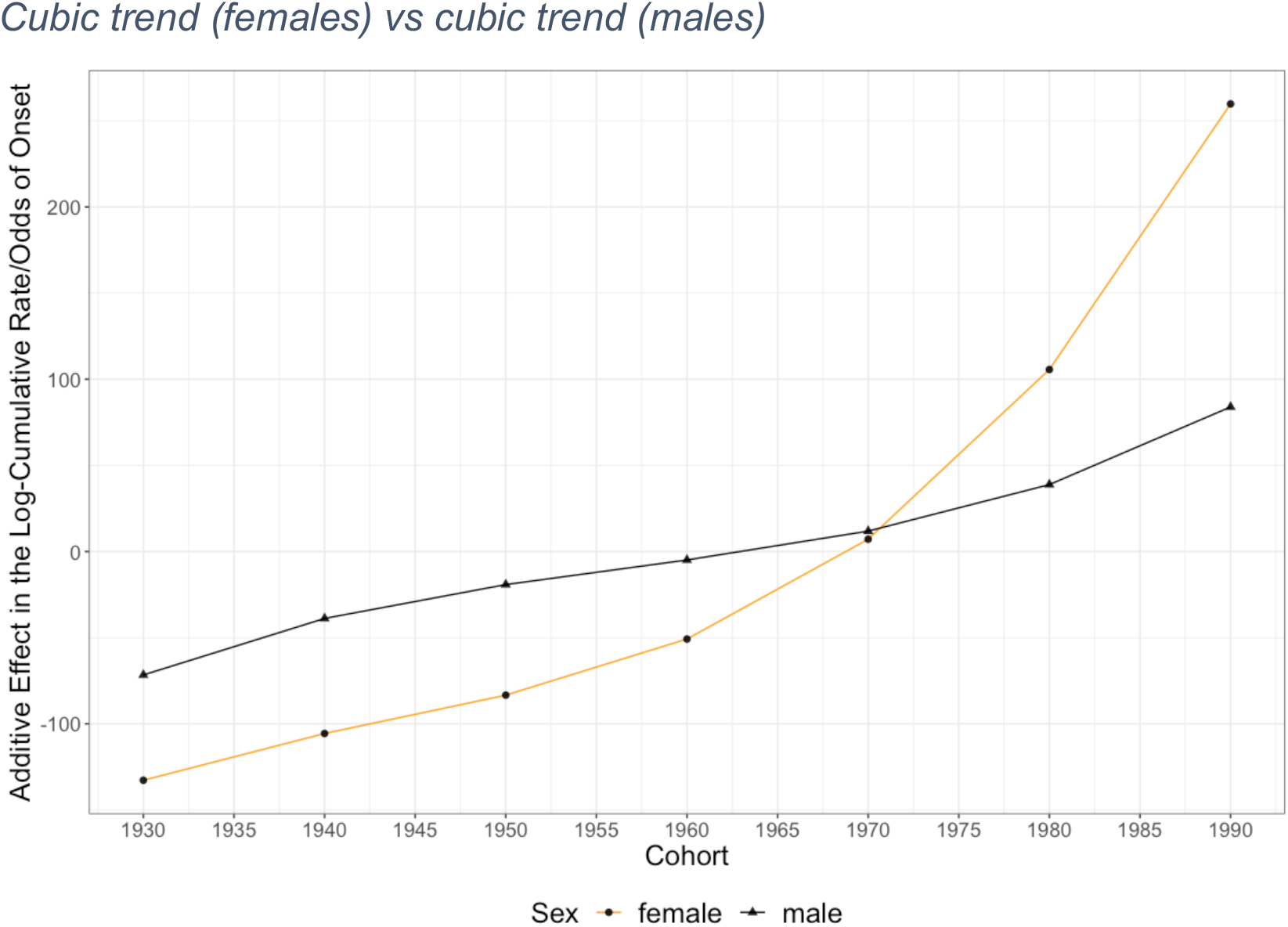
Temporal Trend of the Baseline Log-Cumulative Rates/Odds of Onset Across Cohorts by Sex.

Figure 6 shows the temporal trend across periods predicted by the orthogonal polynomial trend terms by sex, representing the different trajectories across periods that the baseline log-cumulative rates of onset/odds of onset follow for females and males. Across periods, the temporal trend among females was best described by a linear constant trend in contrast to the trend for males, which was best described by a cubic term. This suggests that the rates of onset among females have increased across periods at a constant rate while rates of onset for males have increased at non-constant rates across periods. Since the best fit models for males and females differ in their structural specification of the temporal trend (i.e., linear trend for females, cubic trend for males), Table 4 compares the coefficient estimates of the temporal trends between females and males for models with the same temporal trend specification (i.e., linear vs linear, quadratic vs quadratic, and cubic vs cubic). In all cases, the magnitude of the coefficients for females is greater than for males, suggesting larger rates of increase of the rates/odds of onset for females across periods than for males.

**Table 4.**
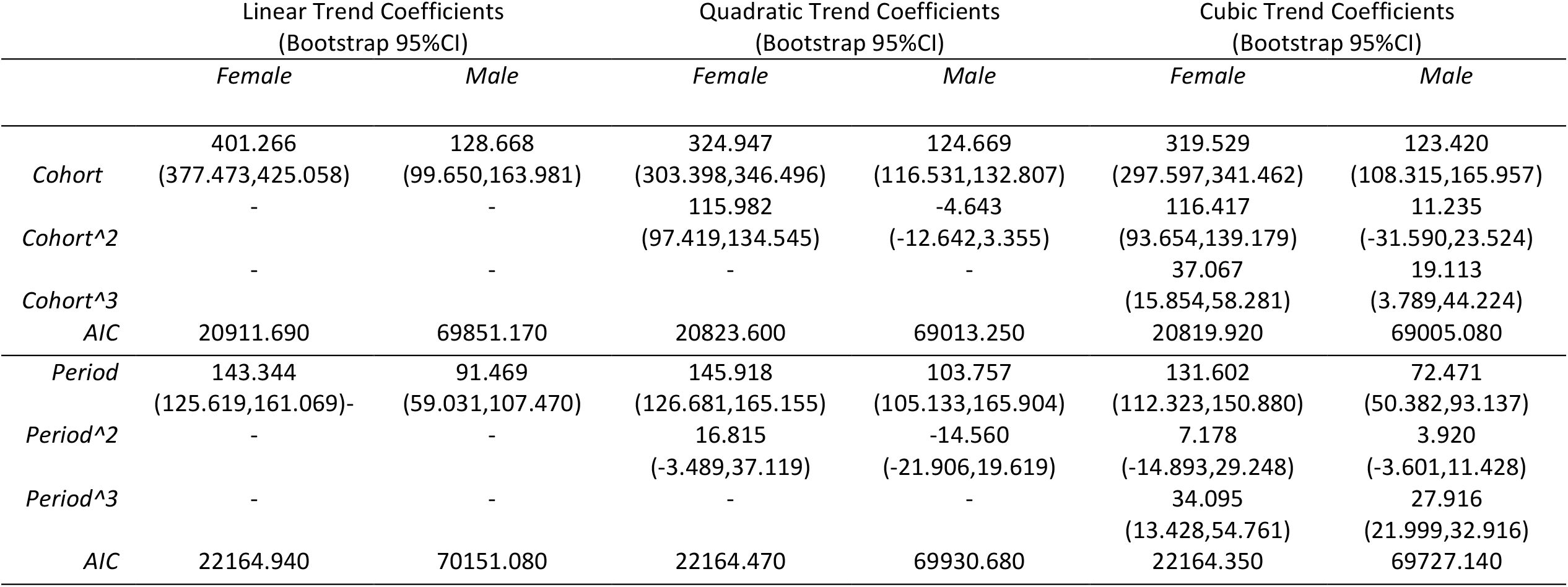
Comparison of Coefficient Estimates and AIC of Same Polynomial Temporal Trend Models by Sex

**Figure 6.**
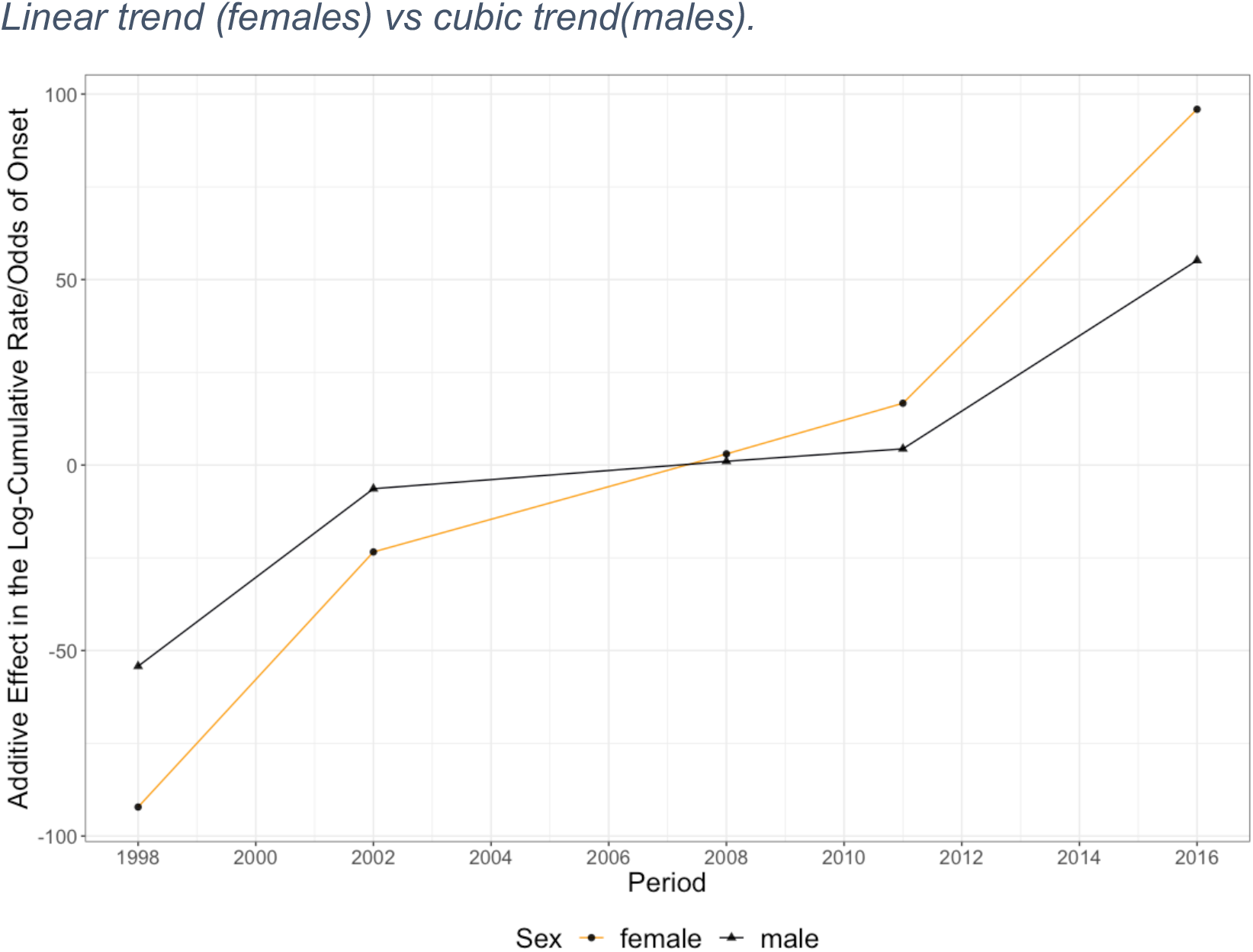
Temporal Trend of the Baseline Log-Cumulative Rates/Odds of Onset Across Periods by Sex.

## Discussion

In this study, we used flexible-parametric time-to-event models to estimate the sex- and age-specific rates of onset of cannabis and their progression across periods and different birth cohorts in Mexico. Our findings support previous estimates on the increasing prevalence of cannabis use in Mexico in the last two decades [29] and confirm the suspicion that the mean age of onset among those who will ever consume cannabis in their lifetime has been reduced for both males and females, reaching similar levels among recent cohorts and periods. We found that age-specific rates of onset of cannabis have increased for both females and males, but females have experienced different growth trends across periods and decennial birth-cohorts than males. In absolute terms, the rates of onset remain different between sexes; rates of onset for males are higher than for females across all periods and cohorts. However, based on our temporal trend estimates, the change on the rates of onset for females accelerated across the most recent cohorts, and exhibit a larger increase across periods than males. Both findings support the hypothesis of a reduction in the gap of onset of cannabis use between female and male adolescents. If these trends continue, our results suggest that the rate at which females initiate cannabis consumption will continue to increasing among the Mexican population.

Our findings have several implications for drug policy and regulation in Mexico. First, the differences in the rates of onset by age, sex, and their progression across cohorts and periods in the Mexican population can inform interventions addressing prevention and harm reduction and can potentially increase their effectiveness. For example, targeted policies, such as behavior-modifying interventions for adolescents [47,48,49], could make a more efficient use of their resources by acknowledging the increasing rates of onset among female adolescents in their design and implementation (i.e., reframing eligibility criteria, and redesigning screening and demographic-based strategies). Future changes to the regulatory framework in the country will potentially alter patterns of cannabis use and perceptions of harms among the population, and in particular, these changes will put an emphasis on prevention and harm-reduction among adolescents [44,45]. Second, evaluations of the effect of such measures should consider the differences in the rates of onset between subpopulations (age, sex, birth-cohort) in their design and implementation, since the effects will likely differ by these characteristics on the population. Finally, more and better evidence on the use of cannabis in the country can also facilitate the implementation of policies that harness or further explore the potential benefits of the medicinal use of cannabis and non-addictive cannabinoids [46].

Our study has several limitations. First, surveys for years 1998 and 2011 were designed to provide estimates that are representative at the national and regional levels. In contrast, surveys for years 2002, 2008, and 2016, followed a sampling frame that provides estimates at the national and state level. Thus, we restricted our analysis to a national level. Second, reporting of “socially desirable” [24] responses could have been exacerbated in the years in which drug-policy in the country shifted to the so-called “War on Drugs.” By controlling for the temporal trend measured as the period of data collection, we account for potential bias in our estimation due to stricter prohibition and its potential impact on the veracity of the responses in the years after the shift in policy was implemented (2002 on-wards). It is possible that this remedy was partially successful, absent additional meaningful data and evidence to further adjust our estimation. However, the potential bias due to “social-desirability” could only affect our estimates downwards by inducing fewer respondents to confirm the age at which they consumed cannabis for the first time or by reporting that they have not consumed cannabis in their lifetimes at the time of the survey. Under this scenario, our estimates should be considered conservative due to the potential under-reporting of events. Finally, due to the still relatively low prevalence of use of cannabis, there was a considerable proportion of observations that were censored, and there may be a considerable proportion of those censored observations that may never experience the event throughout their lifetimes, but that are indistinguishable from the pool of censored observations. To account for this limitation, we implemented methods that correct for censoring of the data; however, we did not further explore the possibility of accounting for the proportion of people in the study population that would never consume cannabis [1, 23]. This limitation can potentially bias our estimation downwards as well, and our estimates should be interpreted as conservative in the same sense as in the previous limitation.

Despite these limitations, our findings bring unique insights into the progression in the ages of onset of cannabis use among seven different generations of Mexicans and highlight how the gender gap related to cannabis use has decreased between 1998 and 2017. Our framework has the potential to increase the fidelity of studies estimating and projecting health and economic burden in the country related to cannabis. Similar flexible-parametric time-to-event models have been previously used to estimate the age-specific rates of onset of smoking [10,11,14] but have not been previously applied to cannabis. For cannabis, Monshowser et. al [20] estimated rates of onset among adolescents non-parametrically (i.e. based on Kaplan-Meier estimates), however, non-parametric models pose a challenge for simulation and extrapolation of the rates of onset using out-of-population characteristics, thus limiting our ability to estimate economic and health burden on populations through mathematical policy models. This is one strength of our analysis because currently there aren’t any available models that yield estimates of rates of onset at the national level in Mexico that can be used for evaluating long-term costs and effects of prevention and harm-reduction policies. Additionally, by using flexible parametric models we estimated nonlinear patterns of onset of cannabis use, which is a common challenge in studying patterns of substance use where the peak of onset is concentrated in a short window of age (i.e. adolescence and young adulthood) and declines rapidly afterwards as age progresses [20]. Finally, we also characterize the progression in time of the age-specific rates of onset with a higher degree of complexity and detail than what is usually achieved with more standard techniques (e.g., fully parametric, or Kaplan Meier), by estimating non-linear temporal trends of the rates of onset of cannabis use across periods and cohorts.

## Conclusion

Surveys related to patterns of substance use have been used to estimate age-specific rates of onset, which are then used to estimate the effectiveness and cost-effectiveness of different drug policies that focus on prevention and treatment. In this study, we estimated that between 1998 and 2016, the age-specific rates of onset of cannabis use in Mexico have increased, and in recent years, the increase in these rates has accelerated for females of recent generations more rapidly than for males. Rates of onset have increased across all periods and across all birth-cohorts. Although males are the most prone to initiate cannabis consumption overall, the gap between females and males has shortened. We provide estimates of the age-specific rates of onset by sex and their progression across periods and decennial birth cohorts that can be used for estimating the potential benefits of a policy and harm-reduction interventions in Mexico and enhance their design and implementation and potentially increase their efficacy.

## Data Availability

Data is publicly availably and maintained for public access by the Government of Mexico. Modeling and statistical analysis will be made available in a public Github repository once the manuscript is submitted for review by a journal.

https://encuestas.insp.mx/ena/encodat2017.php

