## Supplemental material for "Age-Specific Rates of Onset of Cannabis Use in Mexico"

### Appendix

**Dependent variable:** the logarithm of the cumulative rates of onset of cannabis use or the logarithm of the cumulative odds of onset of cannabis use.

Logarithm of the cumulative rates of onset: $lnH[age, cohort]$
Logarithm of the cumulative odds of onset: $lnO[age, period]$

**Failure time (time to event random variable)**

Age of onset (age)**:** denotes the age of first use of cannabis

**Covariates**

Period: periods of data collection in calendar years (1998,2002,2008,2011,2016)
Cohort: decennial birth-cohorts of respondents in calendar years (1930s, 1940s, 1950s,1960s,1970s,1980s,1990s)

**General Modelling Framework: Flexible Parametric Time to Event Models**

The logarithm of the cumulative rates/odds of onset of cannabis use is modelled additively, where the first term on the right-hand side represents the logarithm of the baseline cumulative rates of onset/cumulative odds of onset of cannabis use, which is modelled as a natural cubic spline, $s(\gamma; \log(age))$, with an arbitrary number of parameters $\gamma$. The subsequent terms represent the orthogonal polynomials of the temporal covariate *z* (period/cohort) of arbitrary degree *d*.

a) For the proportional hazards model with homogeneous covariate we use the complementary log-log transformation of the survival function which is equivalent to modelling the logarithm of the cumulative hazard (log cumulative rates of onset) as the dependent variable

$$g[S(age; z)] = ln[-lnS(age; z)] = lnH[age, z] \quad = \ln H_0[\gamma; \log(age)] + \sum_{k=1}^{d} \beta_k (z)^k$$

$$= s(\gamma; \log(age)) + \sum_{k=1}^{d} \beta_k (z)^k$$

b) By analogy, the proportional odds model with homogeneous covariate uses the complementary log-log transformation of the odds ($\frac{1-S(age;z)}{S(age;z)}$) of the event, which yields the logarithm of the cumulative odds(log odds of onset) as the dependent variable

$$g[S(age; z)] = ln[-lnS(age; z)] = lnO[age, z] \quad = \ln O_0[\gamma; \log(age)] + \sum_{k=1}^{d} \beta_k (z)^k$$

$$= s(\gamma; \log(age)) + \sum_{k=1}^{d} \beta_k (z)^k$$

The choice between the scale of the dependent variable (log cumulative hazard vs log cumulative odds) was based on the overall fit of the models (i.e. AIC). The hazard rates (rates of onset of cannabis) were recovered from the predicted log cumulative hazards/odds by the best fit models.

To correct for multicollinearity of the polynomial terms we use orthogonal polynomials.

$$lnH[age,z] \quad = s(\gamma;\log(age)) + \sum_{k=1}^{d} \beta_k f_k(z)$$

$$lnO[age,z] \quad = s(\gamma;\log(age)) + \sum_{k=1}^{d} \beta_k f_k(z)$$

$where\ f_k(z) is\ a\ polynomial\ of\ degree\ k\ in\ z, and\ where\ f_1, f_2, \dots, f_d\ are\ orthogonal\ functions, i.e$:

$$\sum_{k=1}^{d} f_k(z) f_{k'}(z) = f(x) = \begin{cases} 0, & if\ k \neq k' \\ 1, & k = k' \end{cases}$$

**Best-Fit Models for Females**

The specification of the best model fit by decennial birth-cohort (AIC= 20911.69) included: i) three internal knots on the cubic spline, $s(\gamma; \log(age))$, that models the baseline cumulative rates of onset, and ii) a cubic temporal trend (d=3) . We modeled the outcome as the log of the rates of onset.

$$lnH[age, cohort]_{female} = \ln H_0[\gamma; \log(age)] + \sum_{k=1}^{d=3} \beta_k f_k(cohort) \quad = s(\gamma; \log(age)) + \sum_{k=1}^{d=3} \beta_k f_k(cohort)$$

The specification of the best model fit by period of data collection (AIC= 22164.940) included: i) three internal knots on the natural cubic spline, $s(\gamma; \log(age))$, that models the baseline cumulative odds of onset, and ii) a linear temporal trend (d=1). We modeled the outcome as the log of the cumulative odds of onset; the estimated rates of onset were recovered from the estimated odds of onset.

$$lnO[age, period]_{female} = \ln O_0[\gamma; \log(age)] + \sum_{k=1}^{d=1} \beta_k f_k(period) \quad = s(\gamma; \log(age)) + \sum_{k=1}^{d=1} \beta_k f_k(period)$$

**Best-Fit Models for Males**

The specification of the best fit model by decennial birth-cohort (AIC= 69005.620) included: i) seven internal knots on the cubic spline, $s(\gamma; \log(age))$, that models the baseline cumulative odds of onset, and ii) a cubic temporal trend (d=3). We modeled the outcome as the odds of onset; as above, the estimated rates of onset were recovered from the estimated odds of onset.

$$lnO[age, cohort]_{male} = \ln O_0[\gamma; \log(age)] + \sum_{k=1}^{d=3} \beta_k f_k(cohort) \quad = s(\gamma; \log(age)) + \sum_{k=1}^{d=3} \beta_k f_k(cohort)$$

The specification of the best fit model by period of data collection (AIC= 69727.140) included: i) six internal knots on the natural cubic spline, $s(\gamma; \log(age))$, that models the baseline cumulative odds of onset, and ii) a cubic temporal trend (d=3). We modeled the outcome as the odds of onset; as above, the estimated rates of onset were recovered from the estimated odds of onset.

$$lnO[age, period]_{male} = \ln O_0[\gamma; \log(age)] + \sum_{k=1}^{d=3} \beta_k f_k(period) \quad = s(\gamma; \log(age)) + \sum_{k=1}^{d=3} \beta_k f_k(period)$$

#### Temporal Trends

We tested linear, quadratic and cubic temporal trends by period and cohort for female and male populations. We compare the coefficient estimates $\beta_k,\ for\ k = 1,2,\ldots,d$ for models with same orthogonal polynomial basis by sex (i.e. quadratic model for males vs quadratic model for females), shown in Figures 1

*eFigure 1 Comparison of Temporal Trend Coefficient Estimates (linear, quadratic, cubic) by Sex*

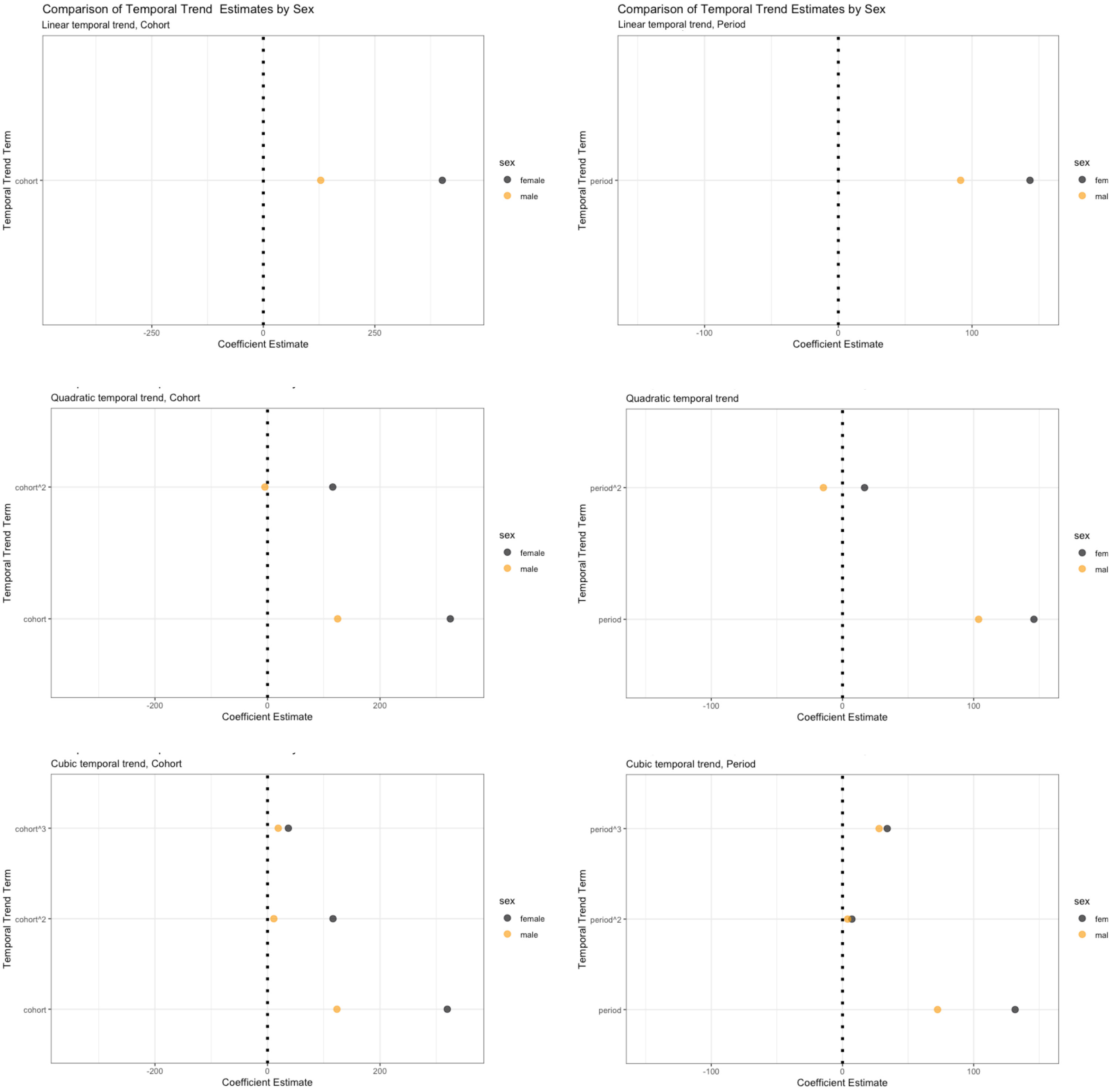

The trajectories  of  the baseline log-cumulative rates/odds of onset predicted by  the same orthogonal functions of period or cohort is compared by sex in Figures 2.

*eFigure 2 Comparison of Trajectories by Orthogonal Polynomial Trend Terms (linear, quadratic, cubic) by Sex*

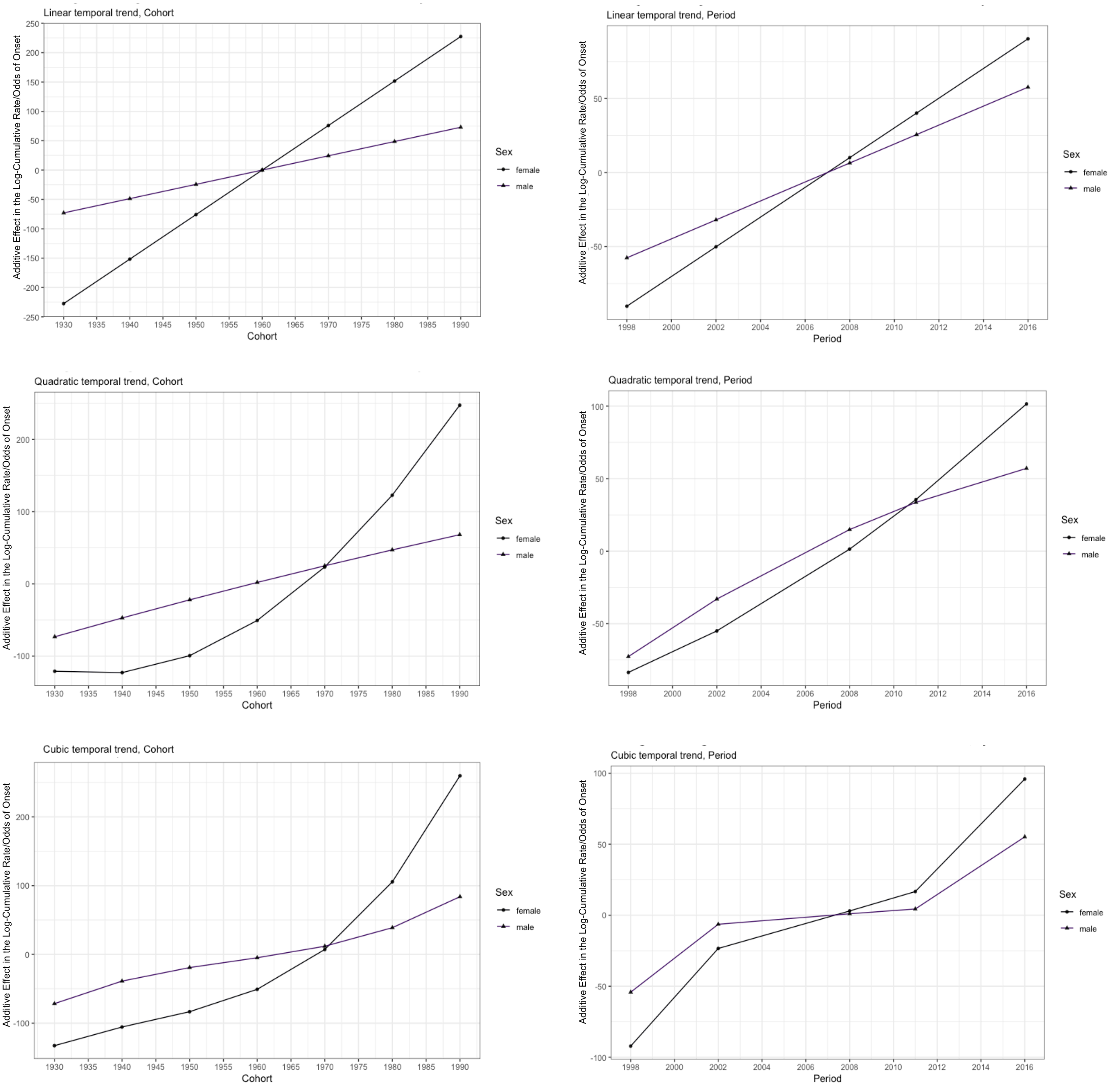

*eTable 1 Log-Rank Test of Hypothesis: Survival time distributions by Sex and Period of Data Collection*

| | N | Observed | Expected | (O-E)^2/E | (O-E)^2/V |
|---|---|---|---|---|---|
| factor(Sex)=Male, factor(Year)=2002 | 403 | 403 | 176.8721 | 289.1004 | 357.5891 |
| factor(Sex)=Male, factor(Year)=2008 | 1583 | 1583 | 1719.039 | 10.7657 | 17.35514 |
| factor(Sex)=Male, factor(Year)=2011 | 730 | 730 | 796.0295 | 5.477055 | 7.564788 |
| factor(Sex)=Male, factor(Year)=2016 | 3050 | 3050 | 3061.394 | 0.042409 | 0.090268 |
| factor(Sex)=Female, factor(Year)=2002 | 54 | 54 | 33.46073 | 12.60766 | 15.42796 |
| factor(Sex)=Female, factor(Year)=2008 | 352 | 352 | 350.9113 | 0.003378 | 0.004342 |
| factor(Sex)=Female, factor(Year)=2011 | 118 | 118 | 132.8531 | 1.660594 | 2.080641 |
| factor(Sex)=Female, factor(Year)=2016 | 928 | 928 | 947.4398 | 0.39887 | 0.563116 |
| | | **Chisquare Statistic** | | **df** | **p** |
| | | 386.8098887 | | 7 | <2e-16 |

*eTable 2 Log-Rank Test of Hypothesis Survival Time Distributions by Sex and Decennial Birth-Cohort*

| | N | Observed | Expected | (O-E)^2/E | (O-E)^2/V |
|---|---|---|---|---|---|
| factor(Sex)=Male, factor(Cohort.dec)=1 | 2 | 2 | 3.539782 | 0.669795 | 0.836887 |
| factor(Sex)=Male, factor(Cohort.dec)=2 | 79 | 79 | 160.7066 | 41.54137 | 54.46745 |
| factor(Sex)=Male, factor(Cohort.dec)=3 | 505 | 505 | 746.6404 | 78.20377 | 111.1677 |
| factor(Sex)=Male, factor(Cohort.dec)=4 | 939 | 939 | 1086.159 | 19.93797 | 29.05147 |
| factor(Sex)=Male, factor(Cohort.dec)=5 | 1335 | 1335 | 1457.834 | 10.34975 | 16.01307 |
| factor(Sex)=Male, factor(Cohort.dec)=6 | 1367 | 1367 | 1329.158 | 1.077415 | 1.636932 |
| factor(Sex)=Male, factor(Cohort.dec)=7 | 1539 | 1539 | 969.2975 | 334.8414 | 481.2862 |
| factor(Sex)=Female, factor(Cohort.dec)=2 | 8 | 8 | 12.21919 | 1.456856 | 1.844232 |
| factor(Sex)=Female, factor(Cohort.dec)=3 | 48 | 48 | 104.8612 | 30.83309 | 41.00014 |
| factor(Sex)=Female, factor(Cohort.dec)=4 | 106 | 106 | 182.631 | 32.15398 | 41.33392 |
| factor(Sex)=Female, factor(Cohort.dec)=5 | 194 | 194 | 274.7454 | 23.73039 | 30.76645 |
| factor(Sex)=Female, factor(Cohort.dec)=6 | 343 | 343 | 404.2087 | 9.268744 | 12.13515 |
| factor(Sex)=Female, factor(Cohort.dec)=7 | 753 | 753 | 485.9994 | 146.6861 | 190.8384 |
| | | **Chisquare Statistic** | | **df** | **p** |
| | | 987.6831172 | | 12 | <2e-16 |

*eFigure 3 Kaplan Meier Age-specific Probabilities of Not Consuming Cannabis for the First Time by Sex and Period of Data Collection (95% CI)*

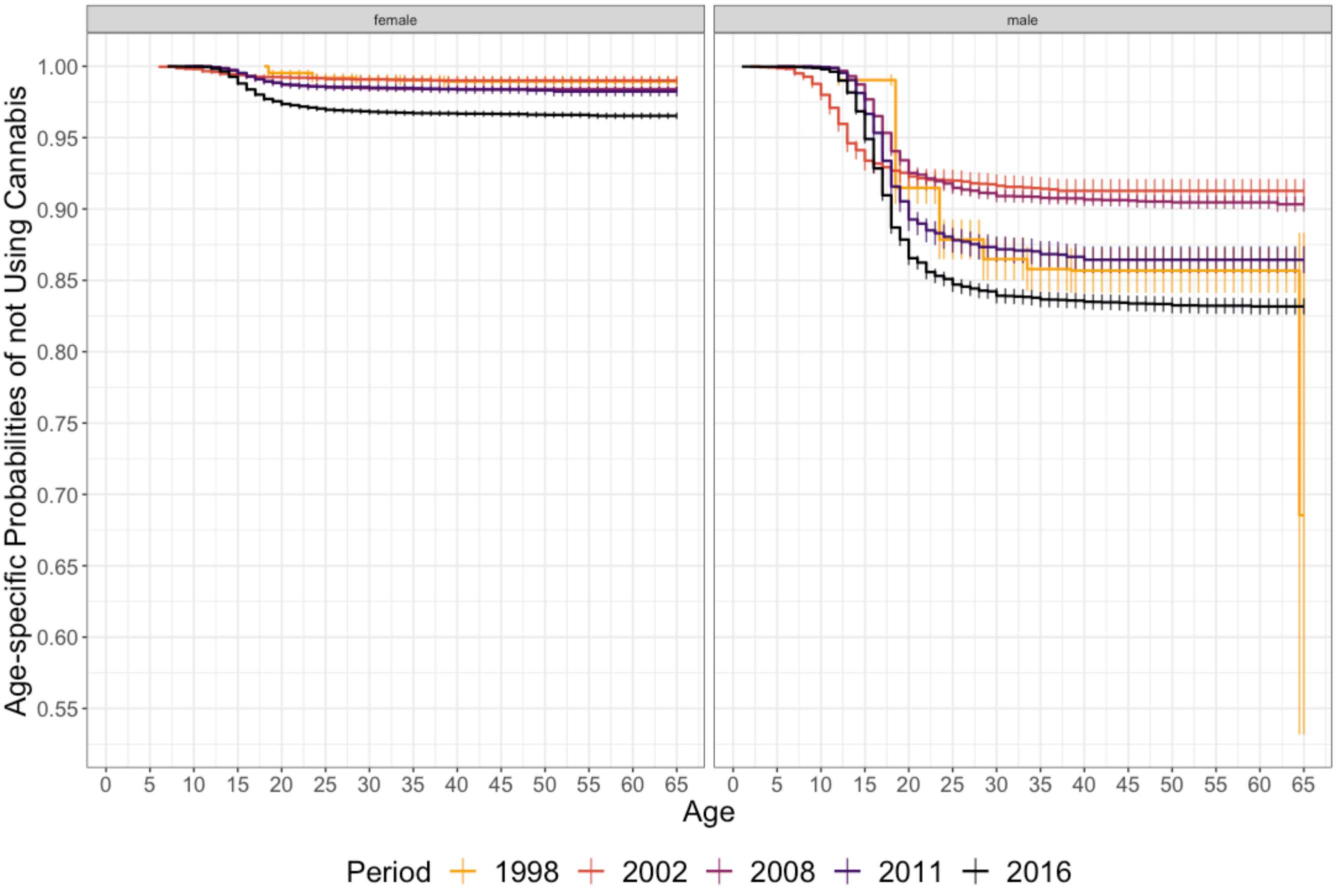

*eFigure 4 Kaplan Meier Age-specific Probabilities of Not Consuming Cannabis for the First Time by Sex and Decennial Birth-Cohort (95% CI)*

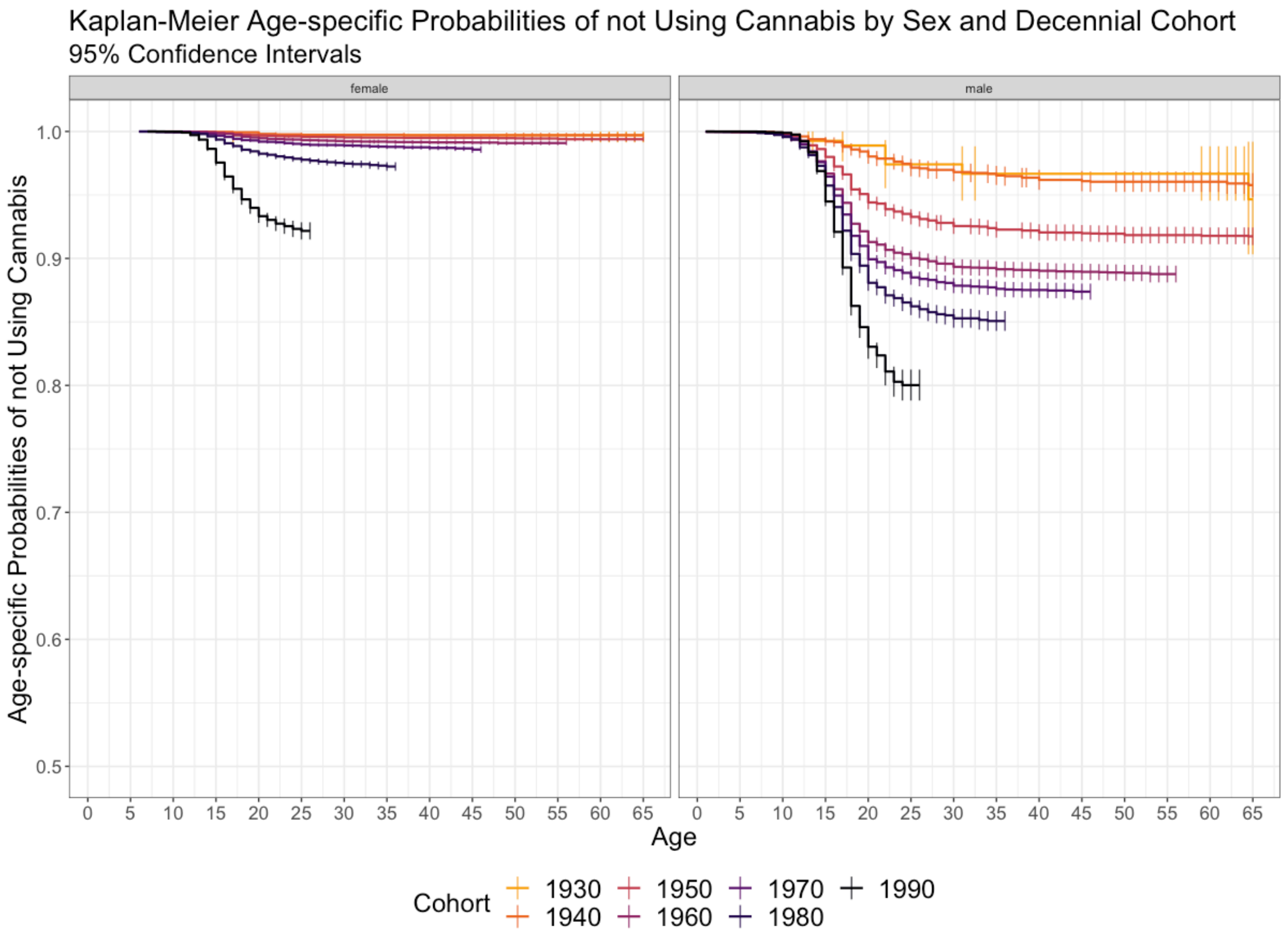
